# Protective characteristics of COVID-19 convalescent and post-vaccination IgG antibodies

**DOI:** 10.1101/2021.11.19.21266547

**Authors:** Melvin E. Klegerman, Tao Peng, Ira Seferovich, Mohammad H. Rahbar, Manouchehr Hessabi, Amirali Tahanan, Audrey Wanger, Carolyn Z. Grimes, Luis Z. Ostrosky-Zeichner, Kent Koster, Jeffrey D. Cirillo, Dinuka Abeydeera, Steve De Lira, David D. McPherson

## Abstract

Soon after commencement of the SARS-CoV-2 disease outbreak of 2019 (COVID-19), it became evident that the receptor-binding domain of the viral spike protein is the target of neutralizing antibodies that comprise a critical element of protective immunity to the virus. This study addresses the relative lack of information regarding actual antibody concentrations in convalescent plasma samples from COVID-19 patients and extends these analyses to post-vaccination samples to estimate protective IgG antibody (Ab) levels. Both sample populations were similar and a protective Ab level of 7.5 µg/ml was determined, based on 95% of the normal distribution of the post-vaccination population. The results of this study have implications for future vaccine development, projection of protective efficacy duration, and understanding of the immune response to SARS-CoV-2 infection.

**One-Sentence Summary:** Using two quantitative immunoassays, we have found similar IgG antibody responses to the SARS-CoV-2 spike protein in populations of COVID-19 survivors and vaccine recipients that indicate a protective antibody concentration.

---

The coronavirus disease outbreak of 2019 (COVID-19), caused by a newly emerging coronavirus, severe acute respiratory syndrome coronavirus 2 (SARS-CoV-2), has infected 253 million people worldwide, resulting in more than 5.1 million deaths (756,000 in the U.S.), nearly two years after initial cases appeared in Wuhan, China, in December 2019 (*1*). The outstanding structural feature of coronaviruses (from which they derive their crown-like designation) is the protruding spikes that mediate attachment of the spherical virions to host cells and subsequent fusion with epithelial cell membranes, required for entry and infection. The spike glycoprotein (SP) that forms these structures is a homotrimer consisting of two subunits, S1 and S2. The S1 protein binds to the angiotensin converting enzyme-2 (ACE-2) receptor on the cell surface through the receptor-binding domain (RBD), while the S2 protein mediates cell membrane fusion (*2*).

It is readily hypothesized, therefore, that the RBD is the target of neutralizing antibodies that comprise the critical element of protective immunity to the virus. This hypothesis is supported by the finding that the RBD is immunodominant and the target of 90% of the neutralizing activity present in SARS-CoV-2 immune sera (*3*). Most compelling is the established 95% protective efficacy of the two mRNA COVID-19 vaccines developed by Pfizer and Moderna, which elicit antibodies specific for the RBD (*4*). Although many studies have focused on characterization of the antibody response to SARS-CoV-2, emphasis has largely been on antibody properties that are most relevant to effective vaccine development, such as the ability of patients to produce high-affinity IgG antibodies specific for the RBD (*2, 5-8*), or production of therapeutic monoclonal antibody formulations (*3, 9-11*). Studies of actual antibody concentrations, binding affinities in terms of equilibrium constants, and non-RBD epitope targeting in convalescent plasma (CP) samples have been largely lacking, however.

We report here the determination of eight different parameters addressing these variables in 21 convalescent plasma samples, using quantitative ELISA methodology (Table 1). We then applied these methods to characterization of 21 plasma samples obtained from individuals who completed the two mRNA vaccination protocols (post-vaccination, PV). Knowing the protective efficacy of these vaccines, which were virtually identical, we were able to estimate protective IgG anti-SP antibody levels and analogous values for two indexes that combined antibody concentrations and binding affinities for both the SP and the RBD.

**Table 1.**
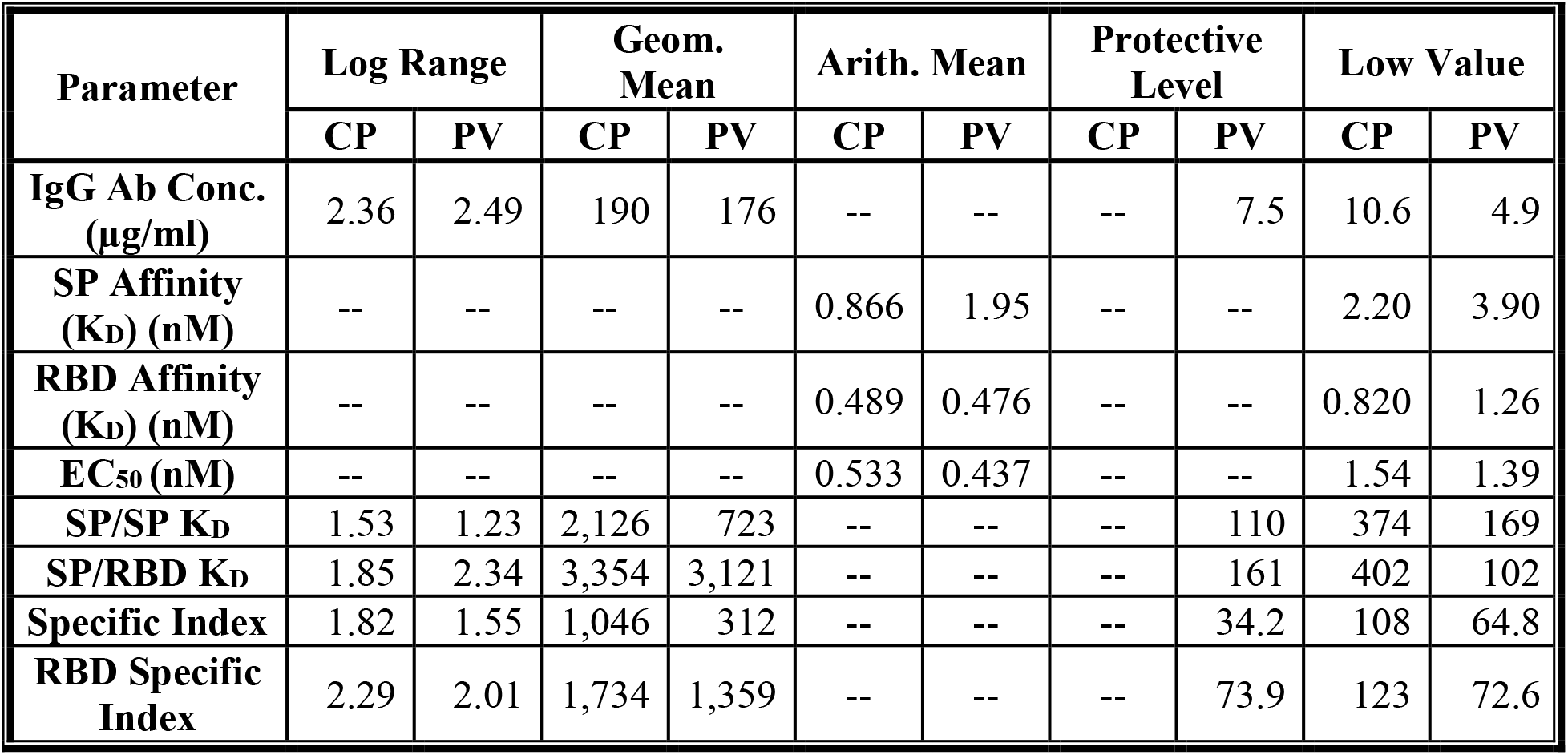
Summary of IgG Antibody Characteristics in Convalescent Plasma (CP) and Post-Vaccination (PV) Samples.

First, we examined Pearson correlations between each pair of four parameters of interest. The results did not reveal any significant correlations between each pair of parameters. The details are shown in Table 2. The overall comparison of means between CP and PV groups for all parameters based on Multivariate analysis of variance (MANOVA) revealed statistically significant differences for at least one of the parameters, (P < 0.002). Further comparisons between the two groups for each parameter by the two-sample t-test also confirmed significant differences between the two groups for SP Affinity, SP/SP (K_D_), and Specific Index between CP and PV groups. The results from both statistical methods are displayed in Table 3. Furthermore, although there was no statistically significant difference between mean IgG antibody concentrations between CP and PV in unadjusted Generalized Linear Model (GLM) (p = 0.35), when adjusted for SP Affinity (KD), there was a significant difference (P = 0.001) between the two groups. Additional results are displayed in Table 4.

**Table 2.** Pearson Correlation Coefficients between IgG antibody characteristics, n=42.

| Parameter | IgG Ab Conc. ($\mu\text{g/ml}$) | SP Affinity ($K_D$) | RBD Affinity ($K_D$) | EC<sub>50</sub> |
| --- | --- | --- | --- | --- |
| **IgG Ab Conc. ($\mu\text{g/ml}$)** |  | 0.37 | 0.15 | 0.06 |
| **SP Affinity ($K_D$)** | 0.37 |  | 0.12 | 0.05 |
| **RBD Affinity ($K_D$)** | 0.15 | 0.12 |  | 0.26 |
| **EC<sub>50</sub>** | 0.06 | 0.05 | 0.26 |  |

**Table 3.** Comparison of the IgG Antibody Characteristics Between Convalescent Plasma (CP) and Post-Vaccination (PV) Groups, (n1=n2=21).

| Parameter | CP<br>(mean) | PV<br>(mean) | T-test | MANOVA analysis |  |
| --- | --- | --- | --- | --- | --- |
|  |  |  | *P* value | *P* value | Overall *P* based on Pillai's trace |
| IgG Ab Conc. (µg/ml) * | 190 | 176 | 0.88 | 0.58 | <0.001 |
| SP Affinity (K<sub>D</sub>) (nM) ** | 0.866 | 1.95 | <0.001 | <0.001 |  |
| RBD Affinity (K<sub>D</sub>) (nM) ** | 0.489 | 0.476 | 0.88 | 0.73 |  |
| EC<sub>50</sub> (nM) ** | 0.533 | 0.437 | 0.34 | 0.16 |  |
| SP/SP (K<sub>D</sub>) * | 2,126 | 723 | <0.001 | <0.001 |  |
| SP/RBD (K<sub>D</sub>) * | 3,354 | 3,121 | 0.87 | 0.80 |  |
| Specific Index * | 1,046 | 312 | 0.002 | 0.002 |  |
| RBD Specific Index * | 1,734 | 1,359 | 0.59 | 0.65 |  |

\* Geometric mean, \*\* Arithmetic mean.

**Table 4.** Mean IgG Antibody Concentration Differences Between Convalescent Plasma (CP) and Post-Vaccination (PV) Groups Before and After Adjustment for SP Affinity (K_D_) or RBD Affinity (K_D_) or EC_50_ in different GLMs.

| Parameter | Unadjusted | Adjusted |  |  |
| --- | --- | --- | --- | --- |
|  | *P* value <sup>1</sup> | *P* value <sup>2</sup> | *P* value <sup>3</sup> | *P* value <sup>4</sup> |
| CP vs. PV | 0.35 | 0.001 | 0.36 | 0.58 |

1: Un-adjusted *P* value for IgG Antibody concentration between CP and PV samples
2: Adjusted *P* value for IgG Antibody concentration between CP and PV samples by SP Affinity ( $K_D$ ) (nM)
3: Adjusted *P* value for IgG Antibody concentration between CP and PV samples by RBD Affinity ( $K_D$ ) (nM)
4: Adjusted *P* value for IgG Antibody concentration between CP and PV samples by $EC_{50}$
\* There is no interaction between IgG Antibody concentration and other parameters

In the recombinant spike protein (rSP) ELISA, the superimposability of normal human plasma spiked with human anti-SP IgG standard (GenScript, Piscataway, NJ), a measure of assay accuracy, is shown in Supplementary Figure 2 (Figure S2). Four normal human plasma samples exhibited antibody concentrations of 28.8 ± 13.0 (SE) ng/ml in the assay, which defines the lower limit of specific COVID-19 antibody response, and recoveries of standard from the pooled normal plasma ranged from 94.5% to 132.5% (mean = 109.7%) from 6.25-100 ng Ab/ml. Antibody standard binding affinity for the spike protein, with correction for post-antibody ELISA incubations (*12*), was found to be 1.46 ± 0.48 nM (K_D_)(SD, n = 29). Binding affinity of ACE-2 Fc for the spike protein was found to be 2.20 ± 0.60 nM (n = 6), which is similar to published values (*2*). Dilutions of both convalescent and post-vaccination plasma samples were superimposable on the respective standard curves (Figures 1A and 1B), indicating that the chimera antibody protein standards are suitable for quantitating anti-SP IgG antibodies in human plasma and serum samples.

**Figure 1.**
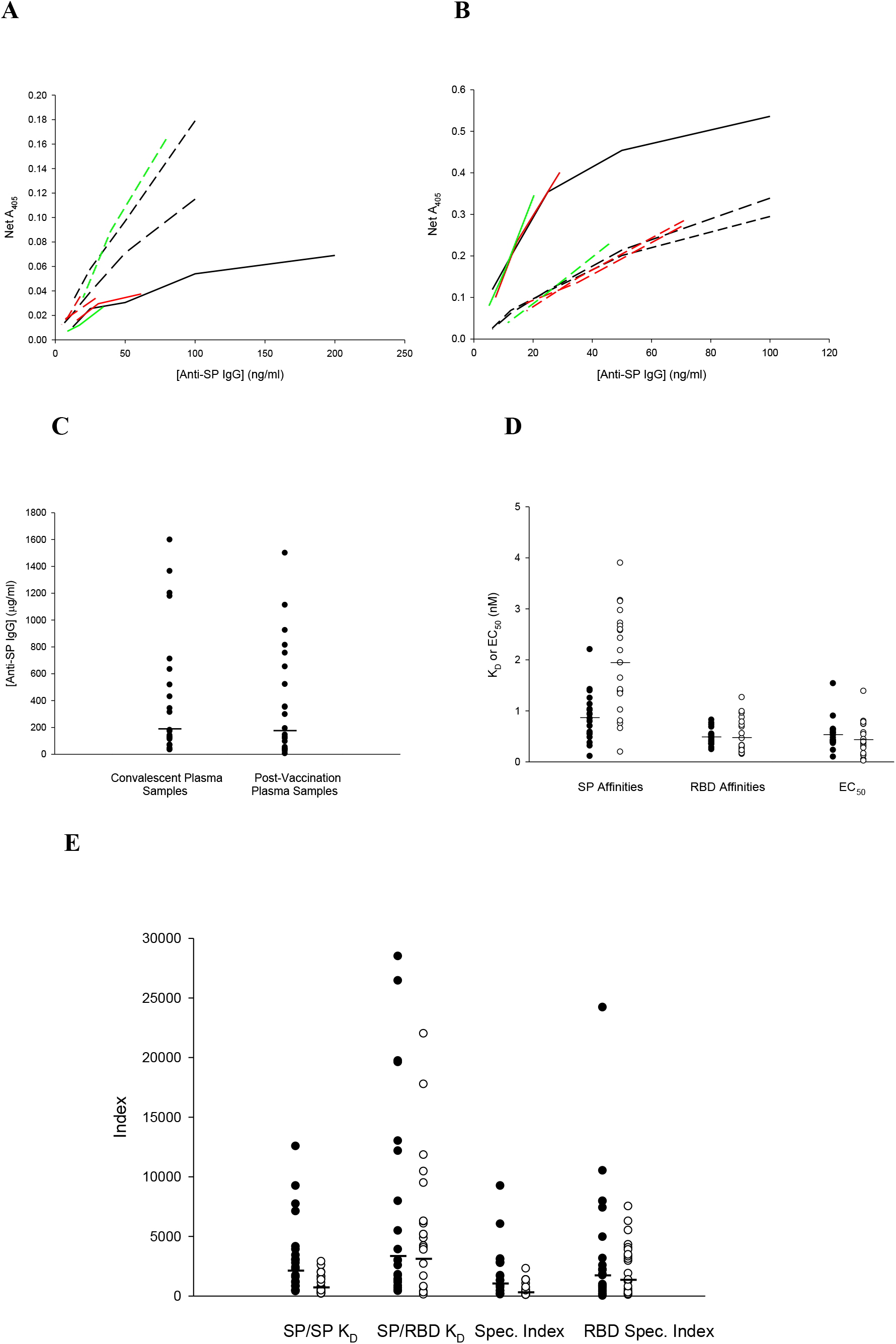
Superimposability of plasma sample dilution responses on anti-SP IgG standard curves. Line plots of five samples relative to three corresponding standard curves, defined by averages of two points at each dilution. Colored lines denote samples assayed with the corresponding standards. **A**. convalescent plasma; **B**. post-vaccination plasma. Point plots of parameter distributions for convalescent and post-vaccination plasma samples. **C**. Anti-SP IgG concentrations. Geometric means are indicated by horizontal lines. **D**. IgG antibody affinities (K_D_) and EC_50_ values. Means are indicated by horizontal lines. **E**. Indexes combining plasma IgG antibody concentrations and antibody binding affinities. Specific indexes are also modified to reflect antibody reactivity with the RBD. Geometric means are indicated by horizontal lines. Convalescent plasma values are denoted by filled circles and post-vaccination plasma values are denoted by open circles in **D** and **E**.

Anti-SP IgG antibody levels for the convalescent plasma population (n = 21) ranged from 33.1 µg/ml to 1.6 mg/ml, while the corresponding values for the post-vaccination plasma population (n = 21) ranged from 4.9 µg/ml to 1.5 mg/ml (Fig. 1C). The geometric means of the two populations were 190 and 176 µg/ml, respectively. The protective antibody plasma concentration, determined as the 5^th^ percentile level of the post-vaccination distribution, is 7.5 µg/ml. SP binding affinities (K_D_) of convalescent plasma samples ranged from 0.1 to 2.2 nM (mean = 0.87 ± 0.47 nM), while the range of RBD binding affinities was much narrower, all being subnanomolar (mean = 0.49 ± 0.15 nM). Spike protein binding affinities of post-vaccination plasma IgG antibodies were lower (p = 0.0001) and somewhat broader (0.2-3.9; 1.95 ± 0.99 nM than those of the convalescent antibody populations. However, the post-vaccination anti-RBD affinities were virtually the same (0.48 ± 0.34 nM; p = 0.877), as were the EC_50_ values, the molar antibody concentration required to produce 50 percent inhibition of a set concentration of ACE-2 binding to the RBD, a putative measure of neutralizing capacity (Figure 1D).

The SP/SP K_D_ index, comprising molar IgG antibody concentration divided by K_D_, reflects the lower antibody affinities of the post-vaccination population, but the values are compacted (Figure 1E). The similarity of the SP/RBD K_D_ values for the two populations reflects the narrow distribution of the RBD affinities, while the lower values of the specific indexes reflect the effect of multiplying the first two indexes by the RBD cross-reactivities, which are mostly less than 100%. Pertinent data for the convalescent patient and post-vaccination recipient plasma antibody populations are summarized in Table 1.

Based on the criteria of log ranges greater than two and low values in the post-vaccination population below the putative protective level, the most useful indexes appear to be the antibody levels themselves, and the two RBD indexes. This may reflect the narrow range of RBD binding affinities, all of which were higher than that of the ACE-2 protein. Comparison of antibody characteristics between the two populations indicates that natural infection with SARS-CoV-2 confers protective levels of humoral immunity on the survivors. Measures of both the quantity and affinity (binding strength) of IgG antibodies for the whole spike protein were greater in the convalescent patient population, possibly reflecting that the mRNA vaccines produce antibodies more specific for the RBD.

None of the parameters significantly correlated with the EC_50_ values. The best correlation for the convalescent plasma samples was found for the specific index (Figure S3A), combining anti-SP Ab concentration, SP affinity and RBD cross-reactivity (r = 0.328, p = 0.171). The best correlation for the post-vaccination samples was found for the RBD affinity (r = 0.362, p = 0.107) (Figure S3B). The Carterra LSA RBD ELISA technology yielded titers for 20 convalescent plasma samples ranging from 20.7 to 1,326 (Geom. Mean = 121). A significant linear correlation was found between the LSA RBD titers and the SP IgG antibody levels (r = 0.464, p = 0.039) (Figure S4), validating the approach by two different methods.

Another approach to validation of the anti-SP IgG concentrations is comparison with previously published studies, all of which expressed antibody levels as some form of titer. Relevant data from 9 studies, including the anti-RBD IgG LSA titers reported in this study, are summarized in Table 5. All but one of these studies were characterized by log titer ranges of 1.81 to 4.70. One very recent study of post-vaccination samples, involving immunoassay-determined levels of anti-SP and anti-RBD IgG and neutralization titers against both pseudovirus and live SARS-CoV-2 virus (*13*), exhibited log ranges for all parameters very close to those found for anti-SP IgG in this study.

**Table 5.**
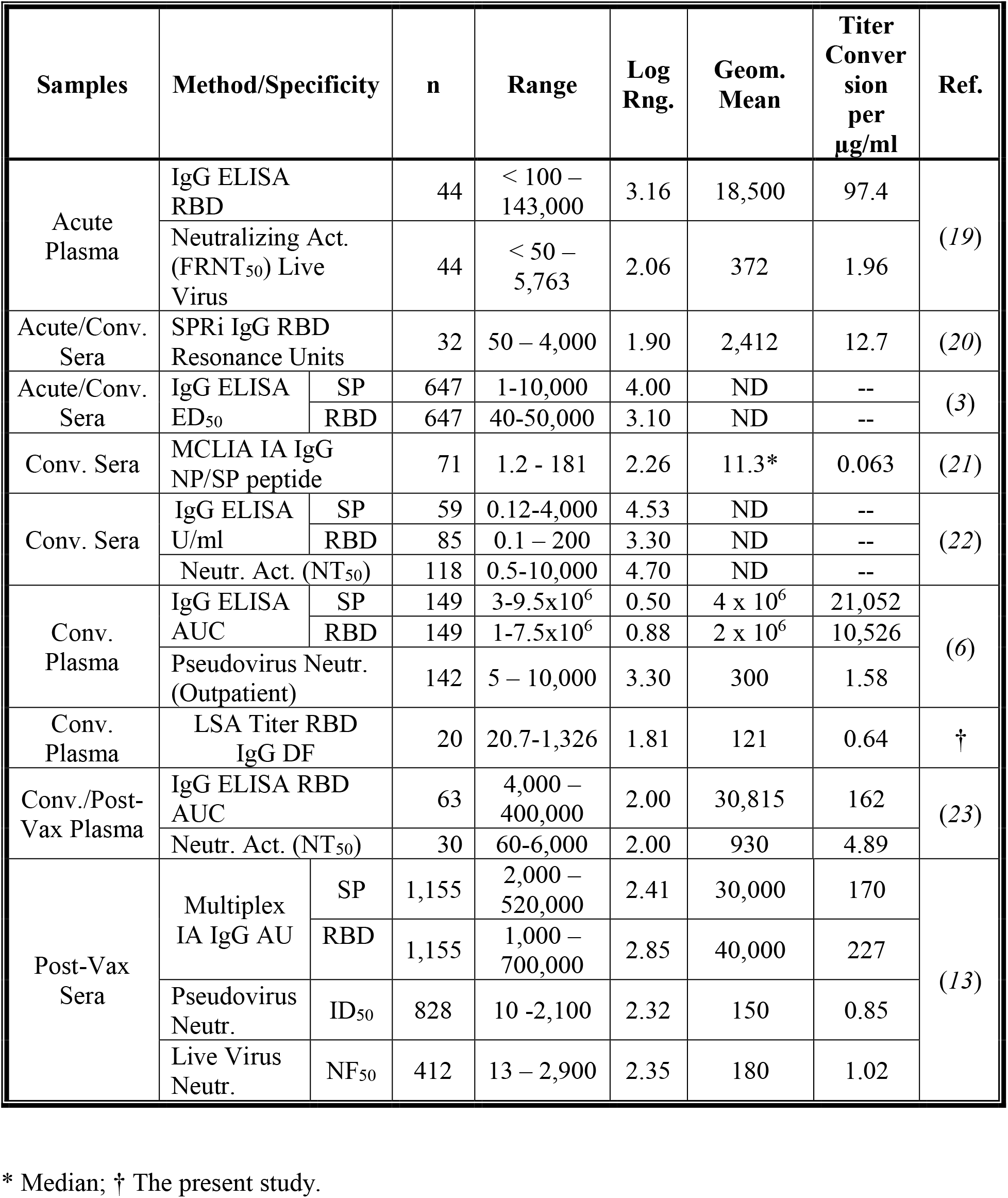
Determination of Antibody Response to SARS-CoV-2 Infection and Vaccination

Factors for conversion of titers to antibody concentrations (µg/ml), based on equivalence of geometric means are included in Table 5. Neutralization titers show the greatest equivalence, with all but one ranging from 0.85 to 1.96. The anti-RBD IgG LSA titers tended to be lower than the corresponding anti-SP IgG levels, which is reflected in a regression slope of 0.300 (Figure S4), and is consistent with the expectation that anti-RBD levels will be lower than anti-SP levels.

In a recent meta-analysis of neutralization data from seven vaccine studies, Khoury *et al*.(*14*) calculated that 50 percent protection against detectable SARS-CoV-1 infection would be afforded by 20.2% of the geometric mean convalescent level of neutralizing antibody. That level for this study is 2.9 μg IgG anti-SP/ml, which is consistent with the 7.5 µg/ml value for 95% protection. Using the conversion factors tabulated in Table 2, the 50% protective values of titers calculated by the Oxford Vaccine Group (*13*) correspond to a minimum of 9.7µg/ml for anti-RBD IgG.

In order to provide a means of standardizing the many COVID-19 antibody assays that have been utilized, the UK National Institute for Biological Standards and Control (NIBSC) has made available panels of pooled convalescent plasma defined by the WHO International Standard in IU/ml (*15*). This standard was utilized in assessing quantitative differences between Moderna (mRNA-1273) vaccine recipients who had COVID-19 breakout infections (n = 36) and those who did not (n = 1,005) (*16*). The geometric mean anti-SP IgG level for infected individuals 28 days after the second vaccine dose was 1,890 IU/ml, while the GM for uninfected individuals was 2,652 IU/ml. The corresponding values for anti-RBD IgG were 2,744 and 3,937 IU/ml, respectively. In view of the results of this study, a much larger difference between the two populations would be expected.

All of these reports emphasized the importance to further vaccine development and refinement of knowing the protective antibody levels in the peripheral circulation. It will also be important to monitoring the decay of protective immunity after vaccination and relating it to the occurrence of breakout infections. It should be emphasized that measuring antibody binding affinity adds value to assessing the robustness of the antibody response, leading to the conclusion in this study that naturally acquired immunity after SARS-CoV-2 infection is comparable to vaccine-induced immunity. It remains to be determined how the former compares to the latter in terms of persistence. These measures will also enable a realistic assessment of immune evasion by viral variants.

Knowing antibody concentrations and target affinities permits a practical understanding of protective immune mechanisms. For instance, based on a SARS-CoV-2 diameter of 100 nm and spike protein surface density of 25 trimers per virion (*17*), a concentration of 7.5 µg anti-SP IgG/ml would provide 12 molecules of antibody per spike protein molecule in a viral saturation of a mucosal surface 10 µm in depth (*18*). With a binding affinity (K_D_) of 1.0 nM, 95 percent of SP molecules will be bound by antibody. With the average binding affinity of 0.48 nM found for anti-RBD IgG in fully vaccinated individuals (Table 1), 99% of SP molecules would be bound by antibody. This calculation provides a conceptual framework that is consistent with the empirical data reported here and the foundation for development of a true protective index based on examination of plasma.

## Data Availability

Small quantities of plasma samples are available to researchers for purposes of reproducing or extending the analysis, subject to materials transfer agreements. Raw data not included in the supplementary materials are available by individual arrangement.

## Author Contributions

Conceptualization: MEK, IS, MHR, MH, AT, JDC, DA, SDL

Methodology: MEK, TP, IS, MHR, MH, AT, DA

Investigation: MEK, TP, IS, MHR, MH, AT

Funding acquisition: MEK, JDC, DDM

Material procurement: MEK, TP, AW, CZG, LZO, KK, JDC

Writing – original draft: MEK, IS, MHR, MH, AT

Writing – review & editing: MEK, TP, IS, MHR, MH, AT, AW, CZG, LZO, KK, JDC, DA, SDL, DDM

## Supplementary Materials

### Materials and Methods

#### Plasma Samples

Convalescent plasma (CP) samples were obtained from the University of Texas Health Science Center at Houston (UTHealth)/Memorial Hermann COVID-19 Convalescent Plasma Program under the direction of Dr. Henry E. Wang. Convalescent plasma (CP) donors previously tested positive for COVID-19, were symptom-free for >14 days, tested negative for COVID-19 antigen prior to donation, and tested positive for CP antibodies. Donor plasma was collected by the Gulf Coast Regional Blood Center, which created a general pool of CP units that were distributed among therapeutic CP programs in the greater Houston area.

Post-vaccination (PV) plasma samples were obtained from Dr. Luis Ostrosky’s laboratory at the Division of Infectious Diseases, Department of Internal Medicine, McGovern School of Medicine of UTHealth. Twenty-one samples collected 15-29 days after administration of the second dose of the Pfizer or Moderna COVID-19 vaccine were studied. Analysis of de-identified convalescent and post-vaccination plasma samples was approved by the UTHealth Committee for Protection of Human Subjects.

#### ELISA for Human IgG Antibodies Specific for the SARS-CoV-2 Spike Protein

This is a direct ELISA in which the antigen is adsorbed directly onto microtiter wells, as we previously described for fibrinogen (*12*). Incubation volumes were 50 µl and all incubations except for the initial coating step were at 37° C. After the blocking step, all incubations were followed by three washes with 0.02 M phosphate-buffered saline, pH 7.4 with 0.05% Tween-20 (PBS-T). Test wells were coated with 5 µg recombinant spike protein (rSP; S1+S2; Creative Diagnostics, Shirley, NY)/ml coating buffer (0.05 M sodium bicarbonate, pH 9.6) overnight at 4° C. Well contents were aspirated and all wells (including background wells for each sample and standard dilution) were blocked for 1 hour with conjugate buffer (1% bovine serum albumin in 0.05 M Tris, pH 8.0, with 0.02% sodium azide). Human anti-SP IgG standards (chimera, GenScript A02038, Piscataway, NJ) (in serial dilutions of 200-12.5 ng/ml PBS-T) and convalescent plasma samples (3 dilutions within the measurable range, previously determined in a screening assay) were incubated in duplicate for 2 hours. All wells were then incubated with 3,000-fold diluted goat anti-human IgG-alkaline phosphatase conjugate (Sigma-Aldrich, St. Louis, MO) for 1 hour. The assay was developed by adding substrate buffer (0.05 M glycine buffer, pH 10.5, with 1.5 mM magnesium chloride) to each well, followed by 4 mg paranitrophenylphosphate (PNPP)/ml substrate buffer (for a total volume of 100 µl/well) and incubating for 15 minutes. The reaction was terminated by adding 50 µl 1 M sodium hydroxide to each well. Plates were read at 405 nm wavelength with a BioTek ELx808 multiwell plate reader.

The OD of background wells was subtracted from test well ODs. The net OD of antibody standard wells was plotted vs. IgG antibody concentration, which obeys a hyperbolic relation. For determination of unknown sample antibody concentrations, the curve fit equation was solved for x. Binding affinities with correction for conjugate incubation perturbation were determined as previously described (*11*). A correction nomogram for this ELISA is reproduced in Figure S1A.

#### ELISA for Human IgG Antibodies Specific for the SARS-CoV-2 Spike Protein Receptor-Binding Domain (RBD)

This is a sandwich ELISA in which a capture antibody, rabbit anti-mouse IgG, is adsorbed onto microtititer wells, followed by antigen capture. Test wells were coated with 2,000X diluted rabbit anti-mouse IgG (Sigma-Aldrich) in coating buffer overnight at 4° C. After blocking, 0.5 µg RSD-mFc (GenScript)/ml PBS-T was added to all wells and incubated for 2 hours. Human anti-SP IgG standards (in serial dilutions of 100-6.25 ng/ml PBS-T) and convalescent plasma samples (3 dilutions within the measurable range, previously determined in a screening assay) were incubated in duplicate for 1 hour. The rest of the ELISA protocol was the same as the rSP ELISA. The ODs of PBS-T only wells run in each assay were subtracted from test well ODs. Calculations of standards and sample antibody concentrations were performed as for the rSP ELISA. A correction nomogram for this ELISA is reproduced in Figure S1B.

#### Determination of Antibody Cross-Reactivity (x-rx) with the RBD

In the spike protein ELISA, binding of ACE-2 to the SP was determined by incubating ACE-2 huFc fusion protein (R&D Systems, Minneapolis, MN) during the primary incubation, instead of human antibody. The anti-human AP conjugate binds to the Fc. Binding affinity was found to be similar to published results (K_D_ = 2.20 ± 0.45 nM, n = 6)(*1*). For determination of x-rx, the A_405_ of replicate wells of 200 ng ACE-2 Fc/ml were measured. One or more dilutions of antibody samples were run with and without admixture of 200 ng ACE-2 Fc/ml. The mean OD of the ACE-2 Fc only wells was subtracted from the OD of each Ab + ACE-2 Fc well and the Ab concentration of these net ODs were calculated (Ab-ACE2). Percent x-rx was calculated as 100-{[(Ab-ACE2} x 100]/Ab alone}.

#### Determination of EC_50_ as a Measure of Antibody Neutralizing Capacity in the RBD ELISA

Three dilutions of each convalescent plasma sample in the range of the ELISA were assayed in the presence of 1 µg recombinant human ACE-2 protein (R&D Systems)/ml, while 1-3 dilutions of the same plasma sample were assayed without the ACE-2 protein in the same assay. Antibody concentration as a percentage of the untreated sample concentration was determined for each dilution in the presence of ACE-1 protein. A linear regression of percent untreated concentration vs. antibody concentration was used to calculate 50% inhibition concentration [(50-a)/b, where % inhibition concentration = b+antibody concentration + a].

#### Indices of Protective Immunity

SP/SP K_D_ is molar concentration of IgG antibody measured in the spike protein ELISA divided by the antibody K_D_ measured in that assay. SP/RBD K_D_ is molar concentration of IgG antibody measured in the spike protein ELISA divided by the antibody K_D_ measured in the RBD ELISA. The specific index is the product of SP/SP K_D_ and SP ACE-2 x-rx as a fraction. The RBD specific index is the product of SP/RBD K_D_ and SP ACE-2 x-rx as a fraction.

#### Carterra LSA RBD ELISA

##### Surface Preparation

An HC200M chip (Carterra PN 4287) was preconditioned via a 2 minute cycling exposure with 50 mM sodium hydroxide (Carterra PN 3638), 500 mM sodium hydroxide, and 10 mM glycine, pH 2.0 (Carterra PN 3640). For goat anti-mouse IgG lawn coupling, the LSA was primed in 25 mM MES, pH 5.5, running buffer + 0.05% Tween-20. The HC200M chip was activated for 8 minutes via cycling exposure of a mixture of 33 mM Sulfo-NHS (Thermo PN 24510), 133 mM EDC (Thermo PN PG82079), 100 mM MES, pH 5.5 (Carterra PN 3625).

Protein coupling was performed by preparing a homogenous anti-murine Fc-specific surface via a 10 minute cycling exposure of 100 µg/ml goat anti-mu Fc specific IgG (Jackson Immunoresearch PN 115-005-071) in 10 mM sodium acetate, pH 4.5 (Carterra PN 3622) + 0.05% Tween-20. The coupling reaction was then quenched by a 6 minute exposure to 1M ethanolamine, pH 8.5 (Carterra PN 3626) + 0.05% Tween-20. Finally, 2 µM RBD-mFc (R&D Systems) was cycled over the anti-mu Fc lawn for 15 minutes.

##### Quantitation (Titer Determination)

Twenty convalescent plasma samples were prepared between two 96 well plates via serial dilutions of each sample, starting with 10X in 1X HBST, pH 7.4 (Carterra PN 3630), and diluting down to 1,280X. Buffer controls and the human anti-SP IgG standards (6.5-830 nM) were also included in each plate. The LSA’s 96 channel printhead sequentially printed the plates on the RBD Lawn. Each plate was cycled via the 96 channel printhead for 30 minutes over the capture surface.

##### Blockade (ACE 2 to RBD-Fc)

After the 30 minute print of each serum plate, the Single Flow Cell (SFC) subsequently injected 588 nM human ACE-2 protein (10-His tag, 85 kDa, R&D Systems) over the chip surface via a 7 minute cycling exposure. The level of ACE-2 to RBD binding was compared between regions that had either serum antibodies bound to RBD or no serum antibodies bound.

### Statistical Analysis

As part of descriptive analyses, we examined the distributions of the IgG antibody characteristics (*e*.*g*., SP Affinity) for subjects in each of the two groups. If the distribution of an IgG antibody characteristic was skewed, we performed log transformation to produce distributions that better represented a normal distribution for subjects in each of the two groups, convalescent plasma (CP) and post-vaccination (PV). We evaluated the linear associations between each pair of IgG antibody parameters using Pearson correlation coefficients or its non-parametric counterpart measure to examine a linearity. In order to control for probability of type I error, first we used multivariate analysis of variance (MANOVA) to compare all means between IgG antibody characteristics. If a significant difference was found, we used t-tests to identify characteristics for which the means between the two groups were statistically different. We used the Generalized Linear Model (GLM) for assessing potential confounding factors or interactions between the study groups and each of the factors associated with IgG antibody concentrations.

---

**Figure S1.**
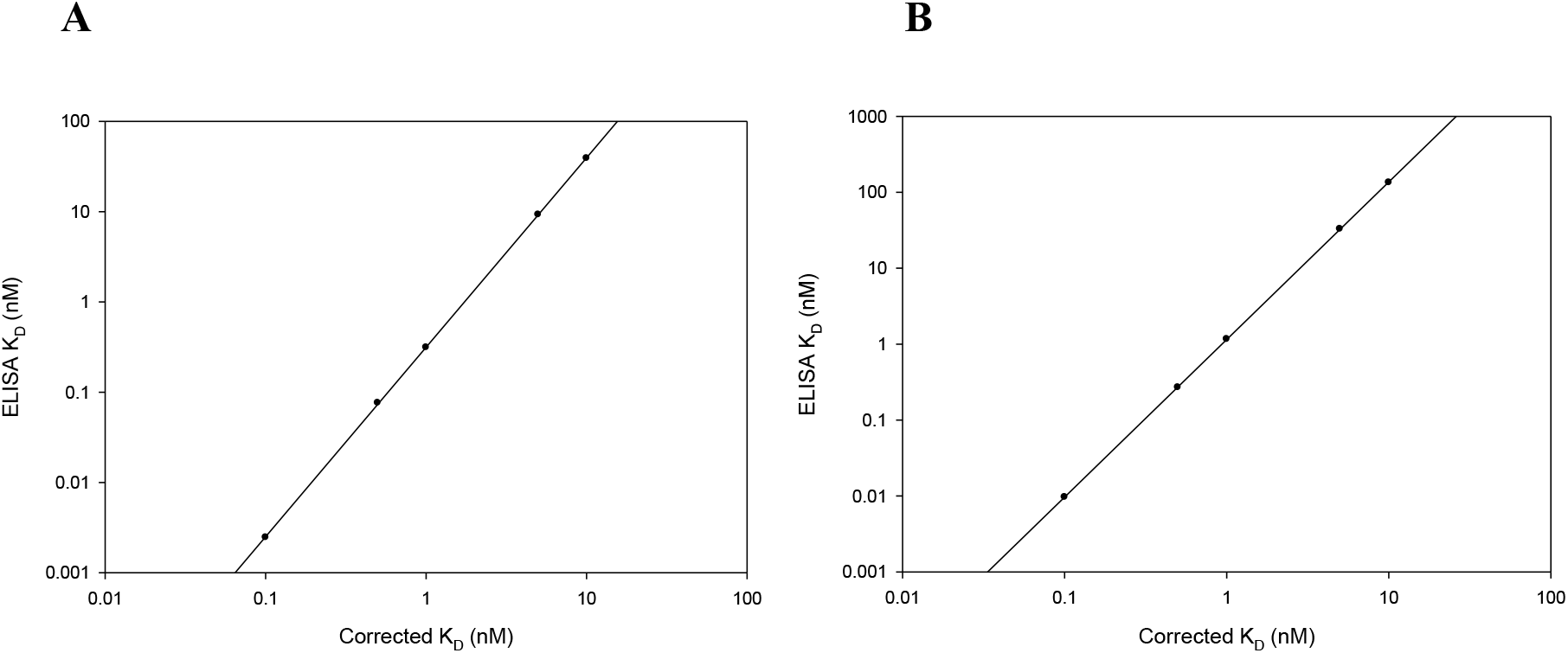
Nomograms for conversion of ELISA apparent dissociation constants (K_D_) to “true” K_D_s, resulting from Underwood correction calculations as described in Ref. 12. **A**. Recombinant spike protein (rSP) ELISA; **B**. Receptor binding domain (RBD) ELISA.

**Figure S2.**
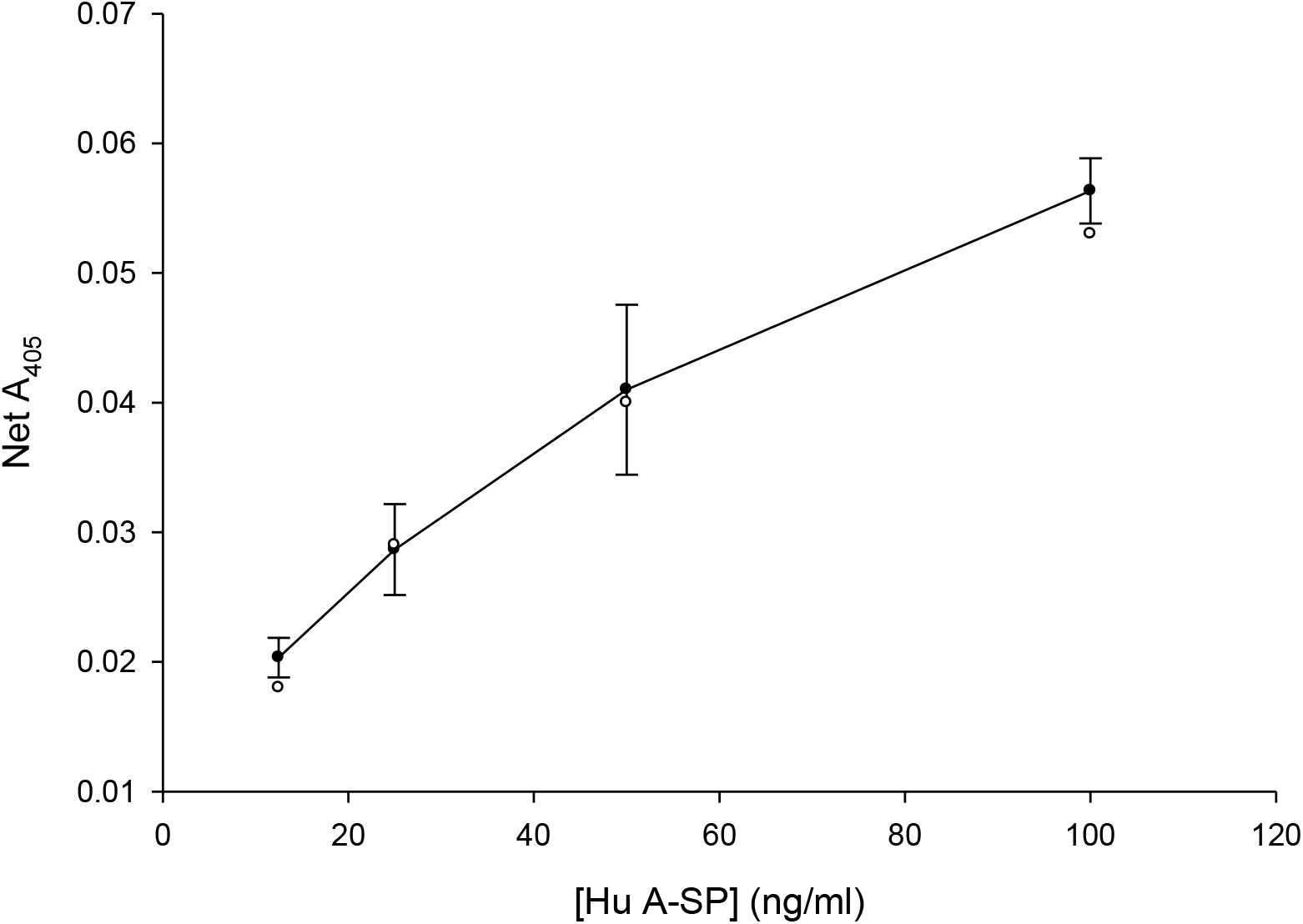
Superimposability of standard-spiked plasma on a composite standard curve in rSP ELISA. Closed circles: standards (mean ± SD, n = 3); open circles: spiked plasma (individual points).

**Figure S3.**
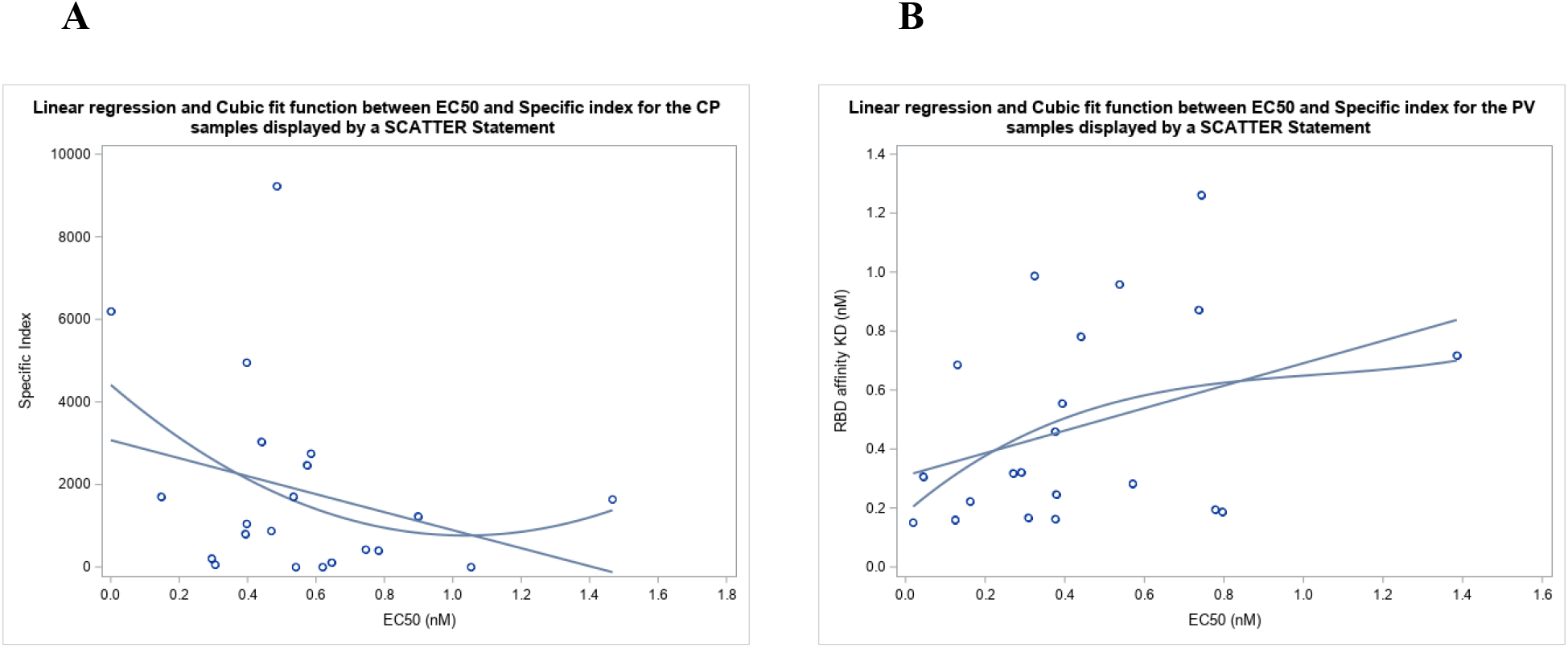
Polynomial (linear and cubit) correlations of EC_50_ vs. calculated parameters. **A**. Specific Index, convalescent plasma (linear fit: r = 0.328; p = 0.171). **B**. RBD affinity (K_D_), post-vaccination plasma (linear fit: 0.362; p = 0.107).

**Figure S4.**
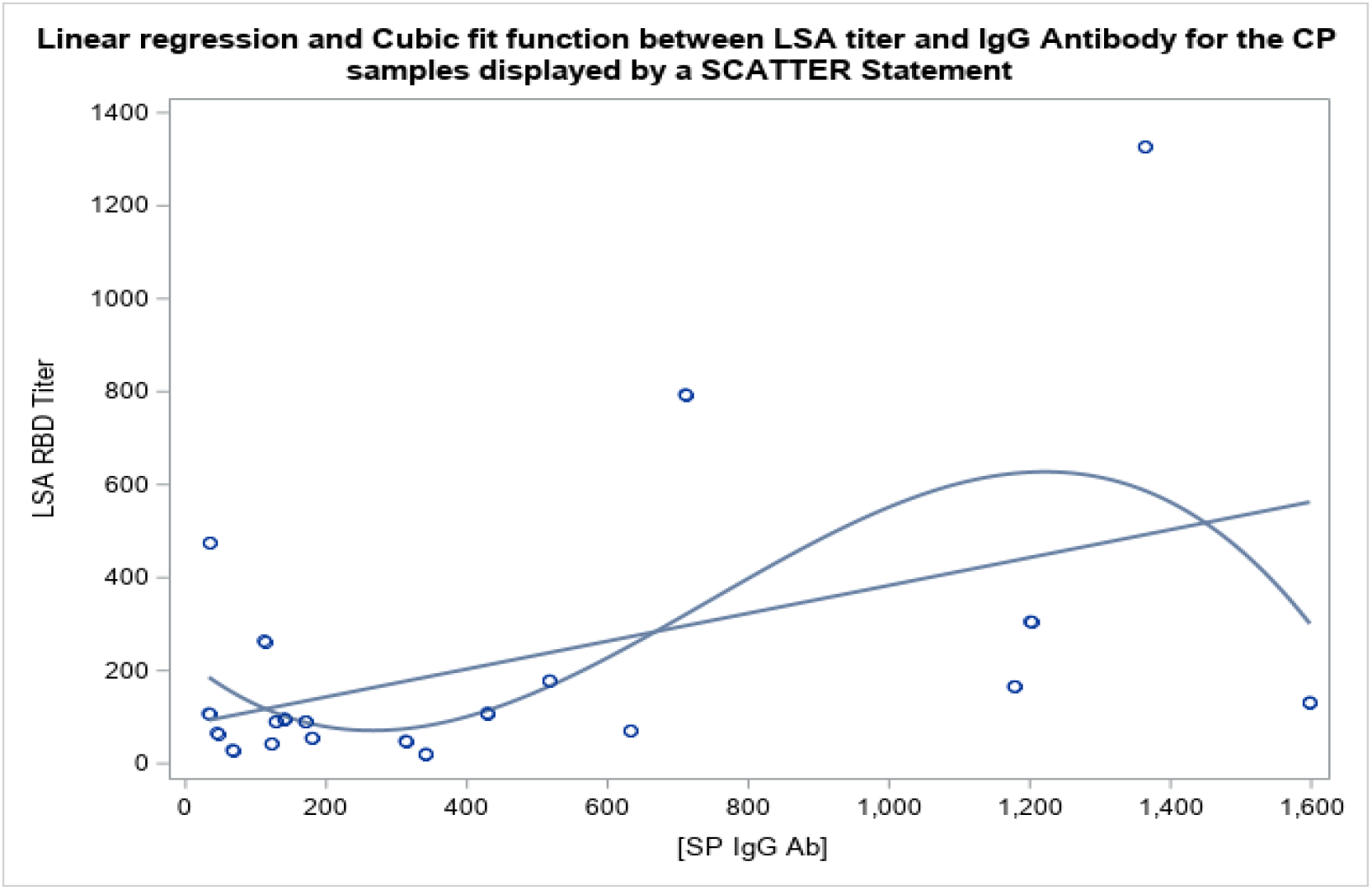
Polynomial (linear and cubit) correlation of convalescent plasma anti-SP IgG concentration vs LSA anti-RBD titer (linear fit: r = 0.464, p = 0.039; y = 0.300 x + 83.5).

